# Safety and efficacy of a modular digital psychotherapy for social anxiety: A randomized controlled trial

**DOI:** 10.1101/2024.07.09.24310160

**Authors:** Mona M. Garvert, Jessica McFadyen, Stuart Linke, Tayla McCloud, Sofie S. Meyer, Sandra Sobanska, Paul B. Sharp, Alex Long, Quentin J. M. Huys, Mandana Ahmadi

## Abstract

**Background:** Social anxiety disorder is a common mental health condition characterized by an intense fear of social situations which can lead to significant impairment in daily life. Cognitive behavioral therapy (CBT) has been recognized as an effective treatment; however, access to therapists is limited and the fear of interacting with therapists can delay treatment seeking. Furthermore, not all individuals respond. Tailoring modular treatments to individual cognitive profiles may improve efficacy. We developed a novel digital adaptation of CBT for social anxiety that is both modular and fully digital without therapist in the loop and implemented it in a smartphone app.

**Objective:** To evaluate the safety, acceptability and efficacy of the new treatment in online participants with symptoms of social anxiety

**Methods:** Two online randomized controlled trials comparing individuals with access to the treatment through the app to waitlist. Participants were recruited online and reported Social Phobia Inventory (SPIN) total scores >= 30. Primary outcomes were safety and efficacy over 6 weeks in 102 women aged 18-35 (RCT #1) and symptom reduction (Social Phobia Inventory total scores) after 8 weeks in 267 men and women aged 18-75 (RCT #2).

**Results:** In RCT #1, active and control arm adverse event frequency and severity was not distinguishable. App acceptability was high. Secondary outcomes suggested greater symptom reduction in the active (-9.83 ± 12.80) than the control arm (-4.13 ± 11.59, t_90_ = -2.23, p_FDR_ = .037, Cohen’s d = 0.47). In RCT #2, there was a higher symptom reduction in the active arm (-12.89 ± 13.87) than the control arm (-7.48 ± 12.24, t_227_ = - 3.13, p_FDR_ = .008, Cohen’s d = 0.42).

**Conclusions:** The online-only, modular social anxiety CBT program appears safe, acceptable and efficacious in an online patient group with self-reported symptoms of social anxiety.

**Trial Registration:** RCT #1: ClinicalTrials.gov NCT05858294, RCT #2: ClinicalTrials.gov NCT05987969

## Introduction

Social anxiety disorder (SAD) is a prevalent and debilitating mental health challenge, affecting a substantial portion of the global population at some point in their lives [1,2]. Characterized by a persistent fear of social situations, SAD can severely limit a person’s ability to engage in everyday activities, from forming personal relationships to navigating work and educational settings. In the long term, SAD can lead to profound social isolation, missed opportunities, and comorbid conditions such as depression, generalized anxiety disorder and substance abuse [1,3,4].

Although by its very nature it often presents late, cognitive-behavioral therapy for social anxiety (CBT-SA) has been established as an effective treatment with moderate to large effects on social anxiety symptoms [5–7]. CBT-SA typically addresses the cognitive processes and behavioral patterns that sustain social anxiety, including negative self-perception in social interactions, self-directed attention, anticipatory and post-event processing of social situations and safety behaviors that, paradoxically, maintain anxiety because they prevent the disconfirmation of negative beliefs [8]. By challenging these patterns, CBT facilitates significant improvements in symptoms, enabling individuals to engage more freely in social situations.

However, traditional CBT-SA faces limitations in accessibility [9] and effectiveness. A significant portion of those suffering from SAD never seek treatment, because interacting with a therapist, a cornerstone of traditional CBT, can be a phobic stimulus [10,11]. Moreover, in many places around the world the availability of trained therapists cannot meet the demand, leading to long wait times and further barriers to accessing care [12]. In response to these challenges, internet-delivered CBT (iCBT), has emerged as promising alternative to traditional therapy [7,13–18]. These programs offer individuals the opportunity to work through therapeutic exercises and techniques at their own pace and in the comfort of their own environment and provide discreet, affordable and immediate support to those in need [19]. iCBT can be supported by a therapist providing guidance remotely throughout the treatment process or be completely self-guided [20–22]. Recent meta-analyses showed that iCBT with and without therapist involvement can be effective in reducing symptoms of SAD [9,22–26].

CBT-SA also does not always work, and can be slow [27]. This may be because standard CBT involves a broad range of interventions aimed at various cognitive and behavioral processes, whereas individual patients may benefit predominantly from a specific subset of these interventions [28,29]. It may hence be possible to further improve treatment efficacy and speed by tailoring interventions to individual cognitive or behavioral profiles [30]. This requires breaking down CBT-SA into distinct, separable modules that target specific cognitive processes or mechanisms selectively [31]. Personalization of treatment may then be achieved by matching interventions to a person’s cognitive profile.

To address accessibility and to work towards a modular targeted therapy, we developed an online-only, modular, iCBT program based on Clark and Wells’ therapy [32]. In it, separate modules target each of the core cognitive components in the standard treatment, including negative beliefs, self-focused attention, rumination, and avoidance behaviors.

Here, we report the findings from two randomized controlled trials investigating the safety, acceptability, and efficacy of this iCBT program in the form of a smartphone app. Overall, the treatment remained safe, acceptable, and effective, significantly improving symptoms of social anxiety in two separate samples compared to a waitlist control group (RCT #1: N = 102, RCT #2: N = 267). This enables the development of mechanistically defined cognitive assessment modules to personalize treatment delivery, and hopefully further improve efficacy.

## Methods

### Objective

We aimed to examine whether the online-only, modular treatment program for social anxiety in the smartphone app was safe and acceptable in the first study, and whether it was efficacious in reducing self-reported symptoms of social anxiety in the second study.

### Ethical Approval

The studies received approval from the Reading Independent Ethics Committee (study reference: AYSATOL). Participants provided informed consent digitally before engaging in any part of the study.

### Clinical Trial Registration

RCT #1 was retrospectively registered on ClinicalTrials.gov after data collection was completed (NCT05858294). For RCT #2, statistical analyses were preregistered on ClinicalTrials.gov before data collection began (NCT05987969).

### Design

We conducted two web-based, unblinded, randomized controlled trials (RCTs). RCT #1 was a six-week parallel-group randomized controlled trial with a four-week intervention and a two-week follow-up. RCT #2 was an eight-week trial with a four-week follow-up. In both trials, participants were randomized 1:1 to the active arm with access to the smartphone app, or the control arm without access to the smartphone app. In RCT #1, participants in the control arm were given access to the smartphone app at week 4, in RCT #2 at week 12.

### Outcomes

RCT #1: Primary outcome measures were safety and acceptability. Secondary outcome measures were symptoms and functioning at week 4 and two weeks post-intervention at week 6.

RCT #2: Primary outcome measures were change in symptoms and daily functioning from baseline to week 8. Secondary outcome measures were safety, efficacy and daily functioning four weeks post-intervention at week 12.

**Safety** was monitored through items in the weekly surveys that asked participants to report any new serious adverse effects experienced in the past week. The Intervention group was asked “Have you experienced any negative effects from using the Alena app? This could be a physical or emotional effect that you believe you have experienced as a result of using the app and/or engaging in the app therapy.” Both groups were asked:

“Have you experienced any new, serious negative health effects in the past week? This includes having to see your GP for a new reason, going to hospital, or being otherwise very unwell in terms of your physical or mental health.” If participants responded positively to either question, they were prompted for additional details and to rate the severity of the event. Any reported events were reviewed by a clinician who determined whether the effect matched criteria for a “Serious Adverse Event”, as defined by the ISO 14155 (A:14).

**Acceptability** was assessed using custom-built questionnaires. Participants were asked how satisfied they were with the app overall (5-point Likert scale from very dissatisfied to very satisfied); how helpful they found the app (5-point Likert scale from very unhelpful to very helpful); how likely they would be to recommend the app (5-point Likert scale from very unlikely to very likely); how easy they found using the app (5-point Likert scale from very difficult to very easy); whether they got to the end of the weekly exercise (yes/no), and what got in the way of completing the exercises, with options provided. Furthermore, adherence to the therapy (monitored by in-app event markers) was monitored through participants’ engagement with the app.

**Symptoms** were measured using the Social Phobia Inventory (SPIN [33]). Designed to evaluate the comprehensive range of symptoms associated with social anxiety – such as fear, avoidance, and physiological reactions – the SPIN includes 17 items, each scored from 0 to 4. This scoring system yields a total possible score ranging from 0 to 68. A score higher than 19 separates individuals with social anxiety from non-anxious controls [33,34]. A decrease of 10 points or more from the baseline SPIN score is considered a reliable indicator of significant improvement in social anxiety, according to the Reliable Change Index provided by The National Collaborating Centre for Mental Health (2018). A score ≤ 19 corresponds to subclinical levels of anxiety.

**Daily functioning** was assessed using the Work and Social Adjustment Scale (WSAS [35]). The WSAS evaluates how much a respondent’s issue affects their ability to perform everyday tasks, including work, managing home responsibilities, and engaging in social and leisure activities. Each activity is rated on a scale ranging from 0 (“Not at all”) to 8 (“Very severely”), with total scores ranging from 0 to 40.

### Eligibility Criteria

Inclusion criteria were as follows:

1. Social Anxiety Symptom Severity: SPIN total score of 30 or higher, indicating a moderate to severe level of social anxiety.
2. Stability on Mental Health Medication: unchanged dose for 8 weeks or more.
3. Age: For RCT #1, participants had to be between 18 and 35 years old. For RCT #2, participants had to be between 18 and 75 years old.
4. Female [RCT #1 only]
5. Technology [RCT #1 only]: smartphone with iOS and internet access. [RCT #2 only]: smartphone with internet access and android or iOS operating system.

Exclusion criteria for both studies were as follows:

1. Alcohol Use: The Alcohol Use Disorders Identification Test for Consumption (AUDIT-C, [36] was used to assess the risk of alcohol dependence. Eligibility required participants to report less than a severe risk level (below eight points out of a possible 12).
2. Recreational Drug Usage: Participants were screened for recreational drug use with the following three questions: Have you used any recreational drugs in the last three months? In the last three months, have you had a strong desire or urge to use recreational drugs at least once a week or more often? In the last three months, has anyone expressed concern about your use of recreational drugs? Eligibility was limited to those reporting minimal to no use (below two points out of a possible 3).
3. Prior Use of Alena App: Participants who had previously used the Alena app were excluded.

### Recruitment

All interactions and data collection occurred online via Prolific, an online recruitment platform. Participants initially underwent a screening process using an online questionnaire that collected information on demographics, lifestyle habits, mental health history, and access to technology. If they passed the screening, they were offered participation in the RCTs.

### Intervention

The treatment consisted of access to the smartphone app. The program was designed in line with the CBT competencies framework [37] and consisted of an introductory module focusing on psychoeducation as well as four targeted modules, each targeting a key mechanism of social anxiety disorder (see Supplementary Material for a detailed program outline):

1. **Introduction:** Serves as an introductory overview, setting the stage for the program and providing insight into the drivers of social anxiety symptoms
2. **Beliefs:** Focuses on conditional beliefs about oneself and others
3. **Attention:** Concentrates on self-awareness and self-focus during social interactions
4. **Avoidance:** Deals with safety behaviors and avoidance patterns
5. **Rumination:** Addresses the tendency to overthink or analyse social interactions after they occur

Each module contained psychoeducational audio lessons and practical worksheets to guide participants through the content (see Supplementary Material). In RCT #2, we updated the visual design and introduced more therapeutic content, including game-like assessments to engage participants further and assess their cognitive and behavioral patterns related to social anxiety. These assessments, lasting between 5 and 15 minutes, were positioned at the start of each module, and completion was required to unlock the rest of the exercises within that module. In RCT #2 participants also had access to the Alena social anxiety community as well as to “Recharge” exercises such as brief meditation and compassion exercises.

### Recharge and Community tabs

Participants in RCT #2 additionally had access to a Recharge and a Community section in the app (Figure 2B). The Recharge section contained mindfulness-based exercises, guided meditations, journaling and self-compassion exercises designed to help participants overcome negative thoughts and feelings related to social situations. For example, participants learned how to observe thoughts without automatically identifying with them, wrote a compassionate letter to the self and were encouraged to celebrate a small win. The anonymous community provided an opportunity to connect with and get support from others by posting in an online chat forum. Participants were suggested to try one Recharge exercise per week and engage with a Community post in some way each week (by liking and/or commenting, and considering posting if they feel comfortable).

**Figure 1.**
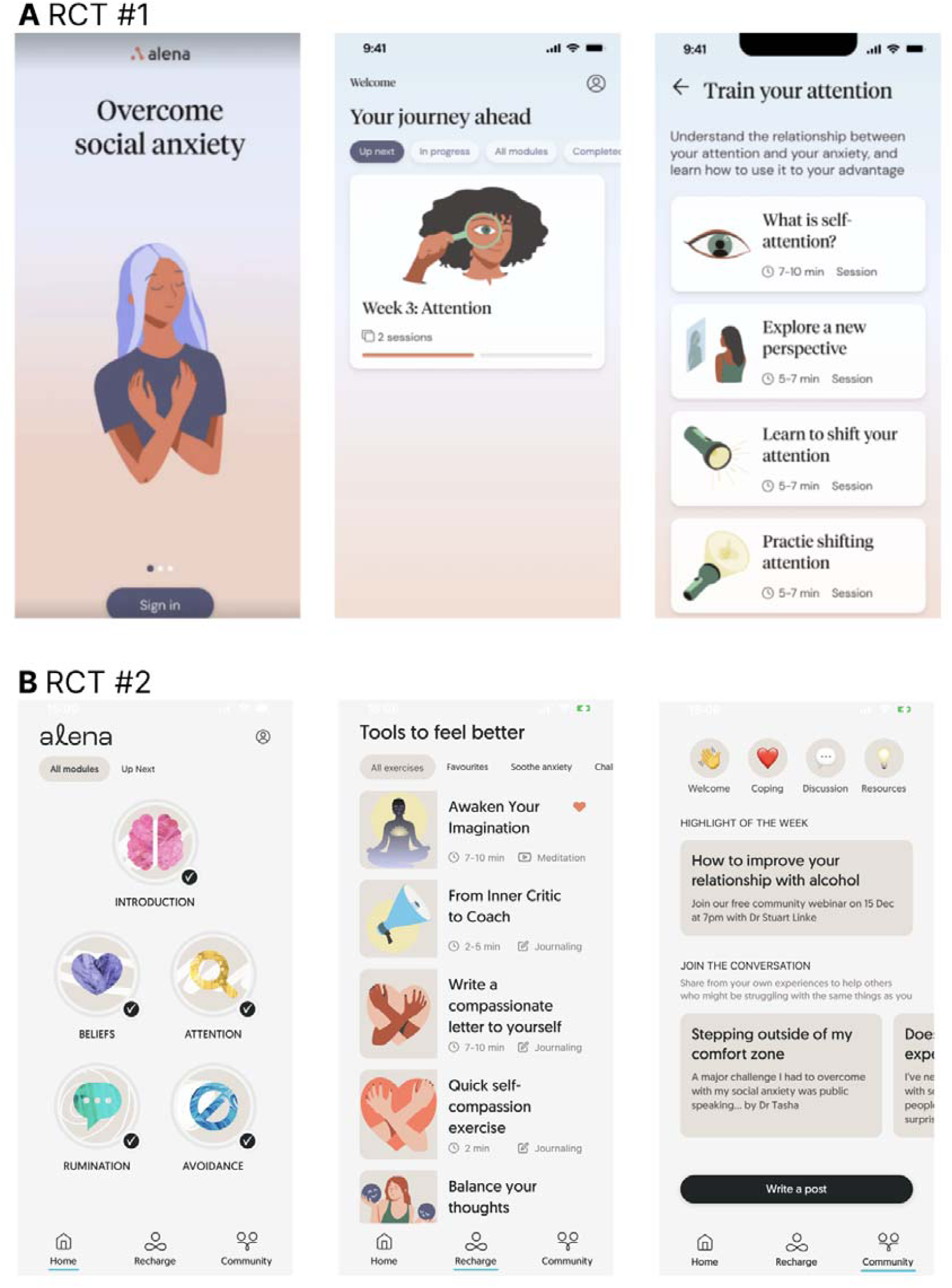
User interface of the apps used in the two RCTs. **A** Interfaces used in RCT #1: (left) the sign-in screen, (middle) program overview with filters for displaying different modules (middle), and (right) a list of exercises for a particular module, e.g., attention. **B** Updated user interface used in RCT #2: (left) the home screen showing each module which could each be tapped to show a list of exercises, (middle) the “Recharge” screen showing a list of exercises not included in the main program but still centred around alleviating social anxiety, and (right) a “Community” screen showing forum posts from members of the Alena community.

**Figure 2.**
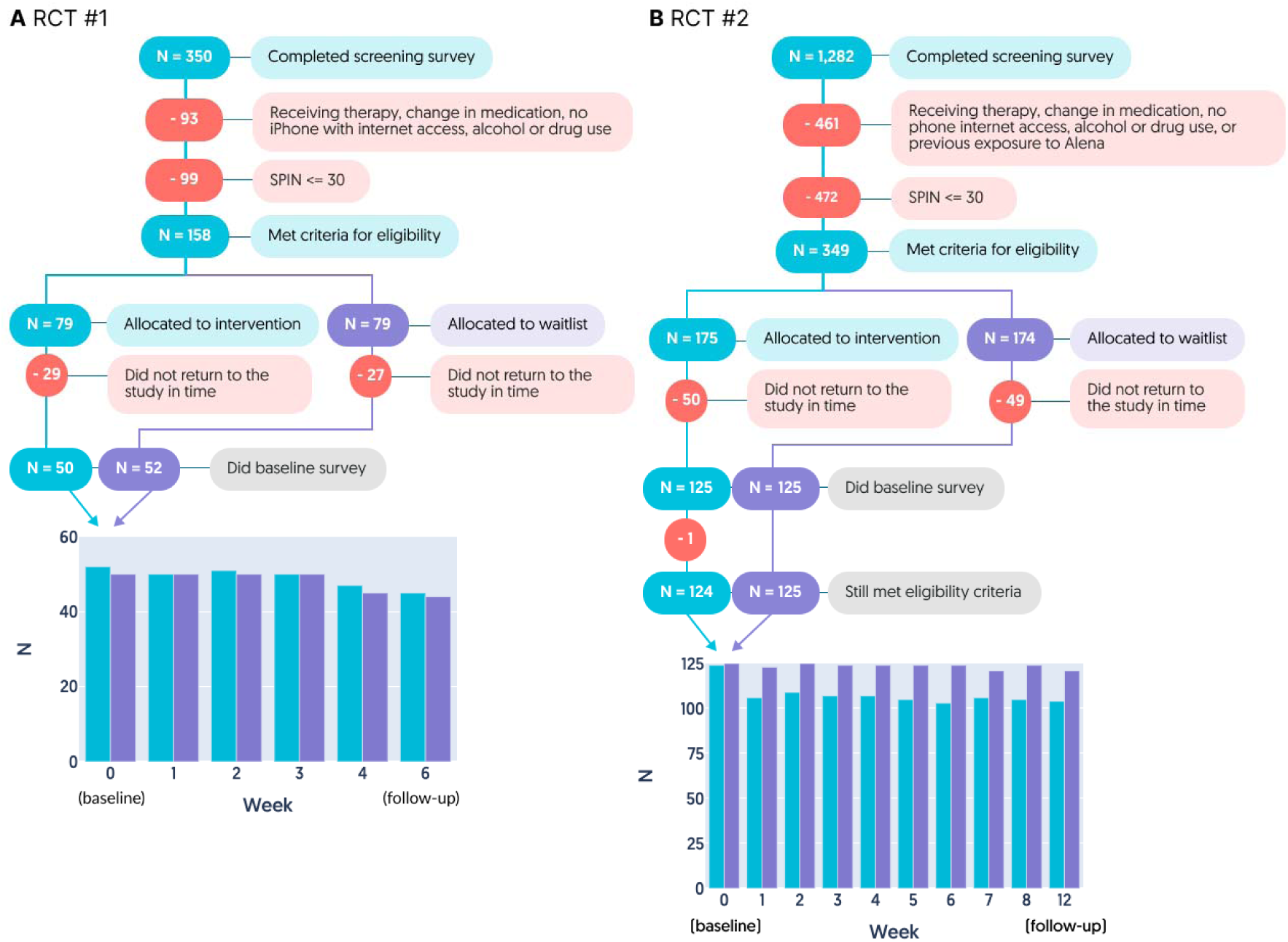
Consort diagram. Flow of participants through the trial for RCT #1 (A) and RCT #2 (B). The number of participants who completed the questionnaires at each time point are visualised at the bottom.

### Program pacing

The exercises, each taking between 1 to 8 minutes to complete, were designed to fit into the user’s daily routine. The app encouraged participants to repeat exercises if needed and to extend their learning outside the app through real-life exposure experiments, supported by in-app exercises that assisted participants with planning and reflecting on these experiments.

To optimise the learning curve and ensure a structured progression through the program, the availability of modules was controlled. In RCT #1, modules were sequentially unlocked each week, while in RCT #2, all modules were accessible from the start, but participants were advised to complete one module every two weeks. To complete all recommended content in the app, participants would have needed to spend between 10 and 20 minutes on the app per week.

### Procedure

Following screening, participants underwent baseline assessments, including SPIN, WSAS, demographics, treatment expectations and previous experience with mental health apps. Participants were then informed of their group assignment. Those in the intervention group received instructions on downloading and using the Alena app, while those in the control group were told they would gain access after a period of four weeks in RCT #1 and twelve weeks in RCT #2. During the intervention or waitlist phase, participants completed the SPIN and WSAS measures every week. Those using the app answered additional questions about their app usage. After the intervention phase, app access was withdrawn from the initial intervention group. A follow-up survey was conducted two weeks later in RCT #1 and four weeks later in RCT #2 to assess short-term maintenance and collect final participant feedback.

### Compensation

The engagement with the app and the therapeutic content itself was not incentivised. However, participants received £1 for their involvement in the screening process. Furthermore, all participants in both RCTs were compensated £5 per survey independent of engagement with the app or therapy.

### Power calculations

Based on a G*Power analysis for the difference between the groups (two-sided t-test), with an estimated medium effect size (d) of 0.6, an alpha level of 0.05, and 80% power, the required sample size is 45 participants per group. Considering a 10% likelihood of participant dropout, we increased our target sample size to 50 participants per group.

Sample sizes for RCT#2 were based on effect size estimates from RCT #1 (Cohen’s d of 0.47 after 4 weeks). To detect effect sizes of 0.47 with an alpha level of 0.05, and a power of 95%, a sample size of 119 participants per group is required. Considering the likelihood of participant dropout, we increased our target sample size to 125 participants per group.

## Statistical Analyses

### Comparison of baseline characteristics between the groups

We conducted Bayesian analyses in JASP (0.18.3) to assess evidence for a null hypothesis that both groups were the same (BF01). Bayesian analysis provides a probabilistic framework that allows for direct comparisons of hypotheses. Specifically, unlike traditional frequentist methods, Bayesian analysis can quantify the strength of evidence for both the null and alternative hypotheses. If BF01 ≥ 3, this indicates evidence for the null hypothesis, whereas a value < 1 indicates evidence for the alternative hypothesis (that the groups are different). A value between 1 and 3 indicates insufficient evidence for either hypothesis. For continuous variables, we implemented Bayesian independent samples t-tests (Gronau et al., 2020), and for categorical or binary variables, we implemented Bayesian contingency tables using an independent multinomial sampling method (groups fixed (Gunel & Dickey, 1974).

### Safety

We compared the number of adverse events between groups using a χ²-test.

### Acceptability

We characterized acceptability descriptively. Dropout rates were also compared using a χ²-test.

### Efficacy and daily functioning

Planned intention-to-treat analyses were performed to compare SPIN (efficacy) and WSAS (daily functioning) scores across groups. Specifically, we used independent t-tests to compare mean changes in scores from baseline to the end of the intervention period, and from the end of the intervention period to the follow-up, between the intervention and waitlist control groups. We used the Benjamini-Hochberg method of False Discovery Rate (FDR) correction to correct for multiple comparisons. Chi-squared tests were used to analyze categorical outcomes across groups, such as the proportion of participants who achieved a clinically significant improvement in social anxiety symptoms (defined as a reduction of ≥10 points in SPIN scores) and those who reached subclinical levels of symptoms (SPIN ≤ 19).

In an exploratory analysis, linear mixed-effects regression modeling was implemented to evaluate the change in SPIN and WSAS scores over time. The models included fixed effects for age, group, time (week), a quadratic time effect (week^2^) accounting for non-linear change in scores over time such as plateau effects and a group × time interaction to assess differential changes in scores between the intervention and control groups across the study period. In RCT #2, additional fixed effects included sex, a group x sex, and a group x week x sex interaction to examine differential changes in scores by sex.

Furthermore, the model included a random intercept for each participant, accounting for the baseline variability in scores among individuals.

Detailed regression tables including full model outputs, including all coefficients, standard deviations, and p-values are provided in Supplementary Tables 1 and 2.

All analyses were conducted using Python version 3.8.9 and R version 4.4.0

## Results

### Participants

Eligible participants from the screening studies (45.14% in RCT #1 and 27.22% in RCT #2, Figure 2) were invited to participate in the main study. RCT #1 included a total of 102 participants (all female, aged between 18 and 35, 50 in intervention and 52 in control group). RCT #2 included 249 participants in total (64% females, 36% males, aged between 18 and 75). One participant in the intervention group of RCT #2 was excluded from the study due to reporting they no longer had access to a smartphone with Internet access in the baseline assessment, yielding N = 124 in the intervention group and N = 124 in the control group.

The groups in RCT #1 were equivalent on all measures except for age, where the intervention group were 1.66 years older on average (mean age = 29.12 ± 4.07 years) compared to the waitlist control group (mean age = 27.46 ± 4.61years old, Bayes factor BF01 = 0.933, Bayesian independent samples t-tests). The groups in RCT #2 were balanced on all baseline characteristics.

**Table 1.** Baseline characteristics of study participants. Group mean and standard deviation (SD) are shown for continuous variables (e.g., age) and the number of participants (N) and group percentages are shown for categorical (e.g., education) or binary (e.g., any drug use) variables. We conducted Bayesian analyses to assess evidence for a null hypothesis that both groups were the same (BF01). If BF01 ≥ 3, this indicates evidence for the null hypothesis, whereas a value < 1 indicates evidence for the alternative hypothesis (that the groups are different). A value between 1 and 3 indicates insufficient evidence for either hypothesis.

| Characteristic | RCT #1 |  |  | RCT #2 |  |  |
| --- | --- | --- | --- | --- | --- | --- |
|  | Intervention | Waitlist | BF<sub>01</sub> | Intervention | Waitlist | BF<sub>01</sub> |
| Age (years) *mean (SD)* | 29.12 (4.07) | 27.46 (4.61) | 0.933 | 39.37 (10.53) | 38.15 (10.84) | 4.910 |
| SPIN - *mean (SD)* | 43.81 (9.14) | 43.28 (7.59) | 4.575 | 44.54 (8.35) | 43.96 (9.34) | 6.348 |
| WSAS - *mean (SD)* |  |  | 4.087 |  |  | 7.192 |
| Expectations for Alena - *mean (SD)* | 2.37 (0.69) | 2.28 (0.73) | 4.055 | 2.27 (0.7) | 2.24 (0.72) | 6.730 |
| Ethnicity |  |  | 37.312 |  |  | > 100 |
| White - *N (%)* | 43 (82.69%) | 43 (86.0%) |  | 103 (83.06%) | 111 (88.8%) |  |
| Black - *N (%)* | 2 (3.85%) | 2 (4.0%) |  | 6 (4.84%) | 1 (0.8%) |  |
| Asian - *N (%)* | 1 (1.92%) | 3 (6.0%) |  | 6 (4.84%) | 8 (6.4%) |  |
| Other - *N (%)* | 0 (0.0%) | 0 (0.0%) |  | 3 (2.42%) | 2 (1.6%) |  |
| Mixed/Multiple - *N (%)* | 6 (11.54%) | 2 (4.0%) |  | 6 (4.84%) | 3 (2.4%) |  |
| Employment |  |  | > 100 |  |  | > 100 |
| Full-time - *N (%)* | 34 (65.38%) | 32 (64.0%) |  | 66 (53.23%) | 62 (49.6%) |  |
| Part-time - *N (%)* | 8 (15.38%) | 8 (16.0%) |  | 24 (19.35%) | 27 (21.6%) |  |
| Student - *N (%)* | 5 (9.62%) | 7 (14.0%) |  | 7 (5.65%) | 5 (4.0%) |  |
| Retired - *N (%)* | 0 (0.0%) | 0 (0.0%) |  | 2 (1.61%) | 2 (1.6%) |  |
| Unemployed - *N (%)* | 2 (3.85%) | 2 (4.0%) |  | 13 (10.48%) | 15 (12.0%) |  |
| Unable to work - *N (%)* | 1 (1.92%) | 1 (2.0%) |  | 6 (4.84%) | 5 (4.0%) |  |
| Temporarily not working - *N (%)* | 2 (3.85%) | 0 (0.0%) |  | 6 (4.84%) | 9 (7.2%) |  |
| Education |  |  | 14.507 |  |  | > 100 |
| No qualifications - *N (%)* | 0 (0.0%) | 0 (0.0%) |  | 1 (0.81%) | 1 (0.8%) |  |
| GCSE or equivalent - *N (%)* | 2 (3.85%) | 2 (4.0%) |  | 16 (12.9%) | 15 (12.0%) |  |
| A-level or equivalent - *N (%)* | 10 (19.23%) | 17 (34.0%) |  | 18 (14.52%) | 30 (24.0%) |  |
| Apprenticeship, higher-education diploma or equivalent - *N (%)* | 4 (7.69%) | 4 (8.0%) |  | 14 (11.29%) | 8 (6.4%) |  |
| Bachelor's degree or equivalent - *N (%)* | 36 (69.23%) | 27 (54.0%) |  | 49 (39.52%) | 54 (43.2%) |  |
| Postgraduate degree or equivalent - *N (%)* | - | - |  | 24 (19.35%) | 14 (0.0%) |  |
| PhD or equivalent - *N (%)* | - | - |  | 2 (1.61%) | 3 (2.4%) |  |
| Alcohol use - *mean (SD)* | 2.71 (1.71) | 2.48 (1.74) | 3.898 | 2.4 (1.97) | 2.02 (2.16) | 2.685 |
| Any drug use - *N (%)* | 5 (9.62%) | 2 (4.0%) | 4.524 | 3 (2.42%) | 6 (4.8%) | 10.461 |
| Ever had therapy for social anxiety - *N (%)* | 44 (84.62%) | 35 (70.0%) | 1.069 | 53 (42.74%) | 50 (40.0%) | 5.860 |
| On medication - *N (%)* | 12 (23.08%) | 9 (18.0%) | 4.180 | 27 (21.77%) | 27 (21.6%) | 7.688 |
| Used apps for mental health before - *N (%)* | 28 (53.85%) | 22 (44.0%) | 2.534 | 38 (30.65%) | 42 (33.6%) | 6.009 |

### Retention

A linear regression with retention as outcome revealed a significant effect of week (p = .004), but not group (p = .47) for RCT #1, reflecting a decrease in retention over time in both groups. In RCT #2, there was a significant effect of group (p < .001), but not week (p = .43) with better retention for the waitlist control group. The week × group interaction was not significant in either RCT (both p > .1), suggesting retention over time was not affected by group.

### Safety

RCT #1: Three participants allocated to the intervention group (6.0%) and eight participants allocated to the waitlist control group (16%) reported adverse effects at some point during the study (χ^2^ = 1.81; p = .178, Figure 3). In total, fewer negative health effects were reported in the intervention group (intervention: four reports, waitlist: 14 reports, χ^2^ = 5.08; p = .024).

**Figure 3.**
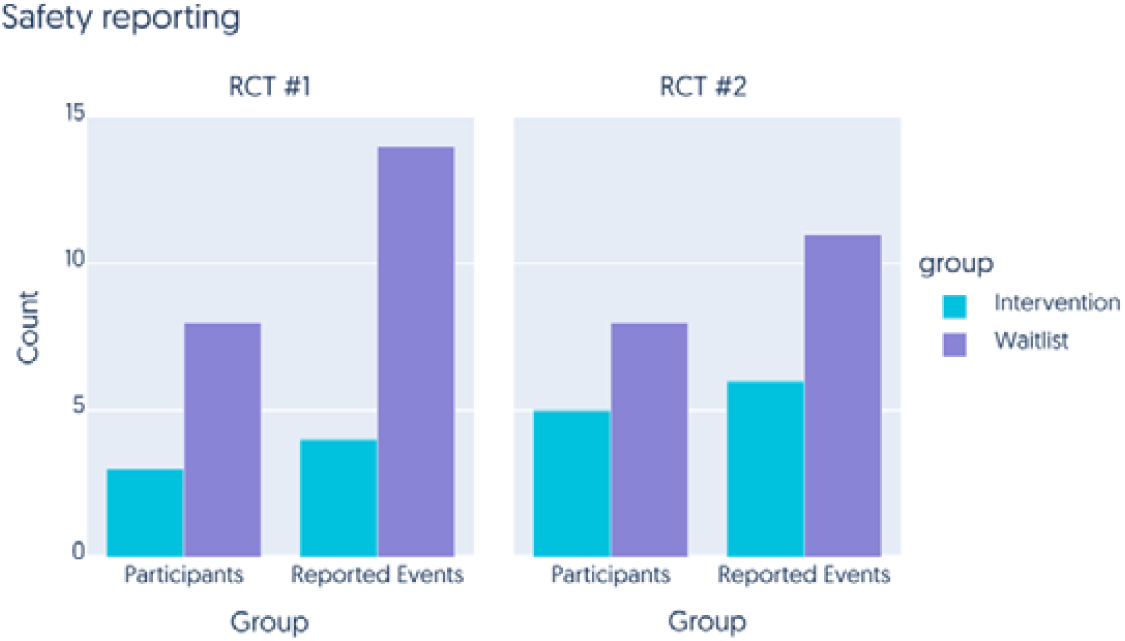
Incidence of adverse health effects. In both RCT #1 (left) and RCT #2 (right), we counted the number of participants reporting serious adverse health effects throughout the intervention/waitlist period, and also the total number of reports made summed across participants, in both the intervention (blue) and waitlist control (purple) groups.

RCT #2: Five participants allocated to the intervention group (4.0%) and eight participants allocated to the waitlist control group (6.4%) reported experiencing adverse effects at some point during the study (χ^2^ = 0.308; p = .579). In total, fewer negative health effects were reported in the intervention group, although this difference was not significant (intervention: six reports, waitlist: 11 reports, χ^2^ = 0.917; p = .338).

Most of the adverse events reported by the intervention group were rated as mild or very mild. Only one adverse event in RCT #1 was rated as serious (“I got covid for the first time and I was hospitalised because of it. I was exhausted and in pain all week.”), but judged to be unrelated to the intervention. The events judged by participants in the intervention group as being related to using the Alena app were mild to moderate in severity, and in line with what would be expected for a psychological therapy, where encountering anxiety-inducing situations in a controlled manner is essential for treatment effectiveness. No severe or very severe negative effects were reported from using the Alena app during the trial.

### Acceptability

To assess user satisfaction and perceived utility, we collected subjective ratings from the intervention group on various aspects of their experience using the app each week. These aspects included overall satisfaction with the app, its perceived helpfulness, the ease of use, and the likelihood of recommending the app to others.

The feedback from participants in both RCT #1 and RCT #2 consistently reflected high levels of acceptability (Figure 4). Participants rated the app highly across all measures, with median ratings reaching 4 out of 5 for satisfaction, helpfulness, and likelihood of recommendation, and median ratings reaching the maximum 5 out of 5 for ease of use. Overall, these findings suggest that the Alena app was highly acceptable to participants.

**Figure 4.**
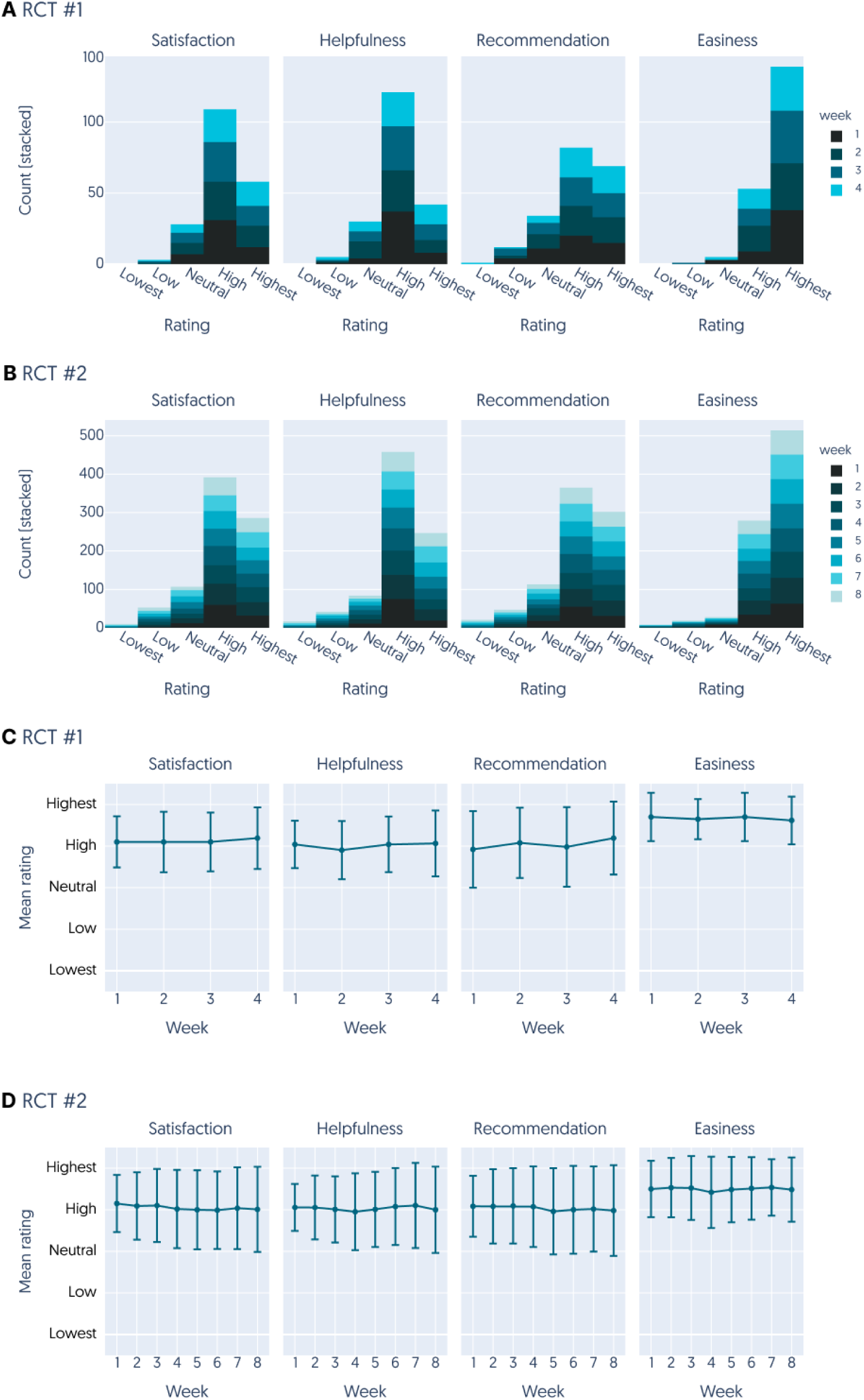
Acceptability ratings of the Alena app. We measured acceptability in four categories: how satisfied participants were with the app, how helpful they found the app, how likely they were to recommend the app, and how easy the app was to use. Response ranged from 1 (lowest) to 5 (highest). Measures were taken each week (see legend for colour scale) in both RCT #1 (**A**) and RCT #2 (**B**). **C** and **D** visualize average ratings across weeks. Error bars denote the standard deviation.

### Therapy adherence

Throughout the intervention period, we tracked how well participants adhered to Alena’s therapy program, monitoring the number of audio lessons listened to and interactive worksheets finished by each participant. Even though participants were not incentivised to adhere to the therapy program (they were only compensated for the time required to complete the weekly surveys), participants in RCT #1 showed a median completion rate of 90.91% (mean = 76.92%, SD = 29.17%, Figure 6). For RCT #2, which featured a longer treatment program, the median completion rate was 84.85% (mean = 73.83%, SD = 27.89%).

**Figure 5.**
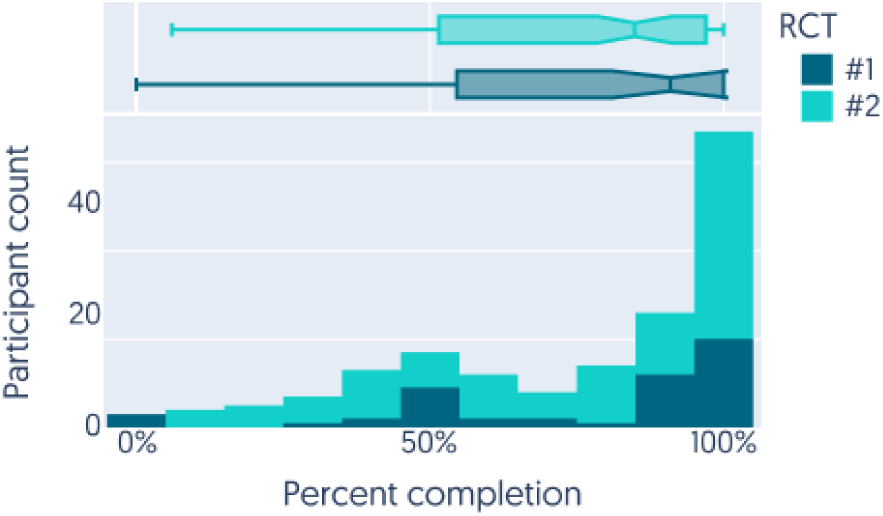
Therapy completion rates. Histogram shows the proportion of exercises completed across participants in the intervention group for RCT #1 (dark blue) and RCT #2 (green). Box-and-whisker plot (top) shows the distribution of therapy completion rates across participants in each RCT, with the median at the notch, the 25th to 75th percentiles represented by the box (i.e., the “interquartile range”), and the “whiskers” of the plot representing each box boundary ± 1.5 × the interquartile range.

**Figure 6.**
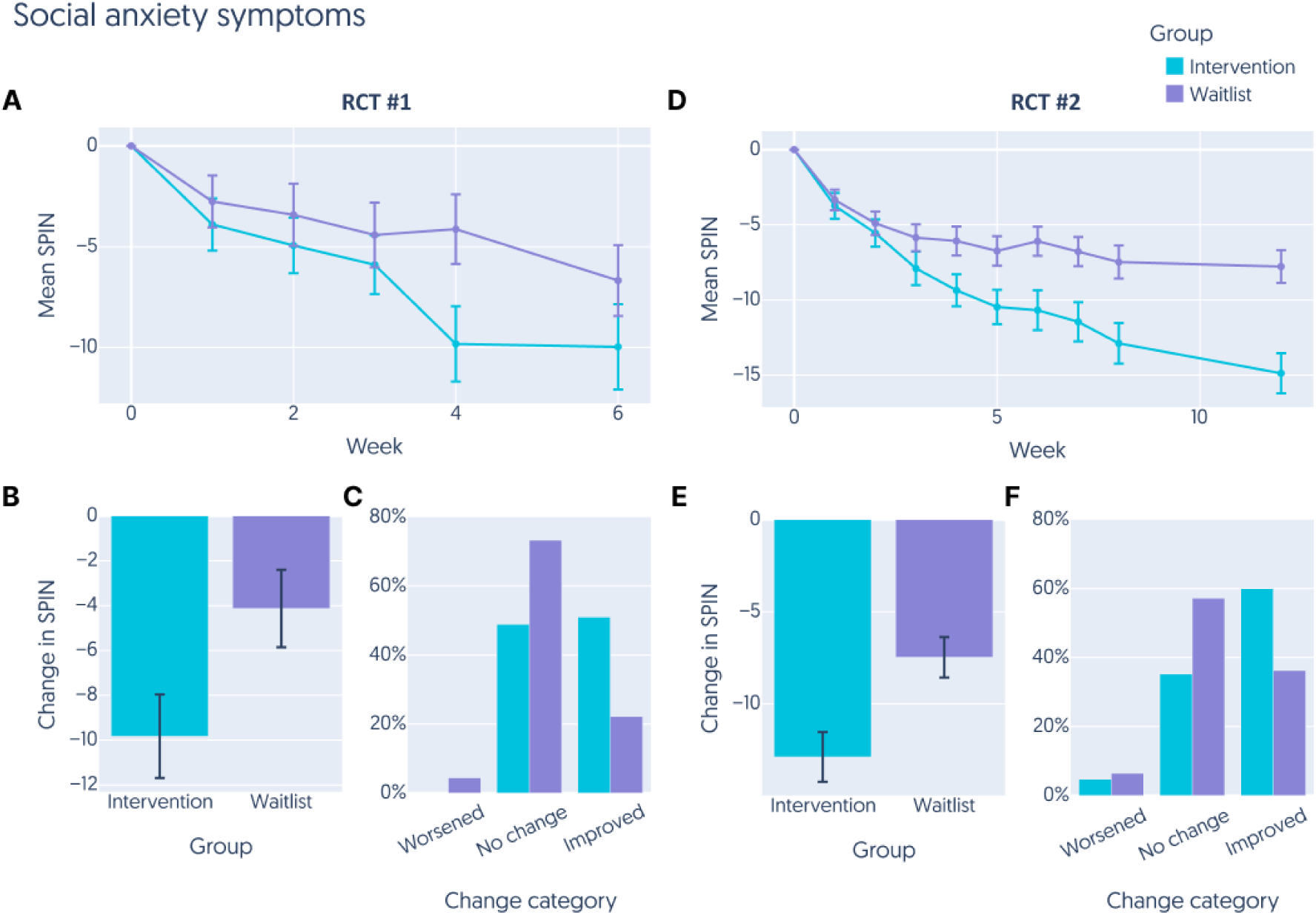
Improvement in social anxiety symptoms over time in both RCTs. **A** The mean SPIN score for participants in RCT #1. Participants in the intervention group are indicated by blue lines, participants in the waitlist control group are indicated by purple lines. SPIN scores were tracked in each week of the study, with the first week (week 0) being the baseline assessment and the last week being the follow-up assessment. All weeks in between were the intervention/waitlist period. **B** Total change in SPIN from week 0 to the final week of the intervention/waitlist period in RCT #1. **C** Proportion of participants in each group whose change in SPIN indicated a worsening of symptoms (increase in SPIN of 10 points or more), an improvement in symptoms (decrease in SPIN of 10 points or more), or no change in RCT #1. **D** The mean SPIN score for participants in RCT #2. Participants in the intervention group are indicated by blue lines, participants in the waitlist control group are indicated by purple lines. SPIN scores were tracked in each week of the study, with the first week (week 0) being the baseline assessment and the last week being the follow-up assessment. All weeks in between were the intervention/waitlist period. **E** Total change in SPIN from week 0 to the final week of the intervention/waitlist period in RCT #2. **F** Proportion of participants in each group whose change in SPIN indicated a worsening of symptoms (increase in SPIN of 10 points or more), an improvement in symptoms (decrease in SPIN of 10 points or more), or no change in RCT #2. Error bars represent the standard error of the mean.

### Efficacy

In both trials, all participants started with a median SPIN score of 43, corresponding to severe symptoms of social anxiety. Symptom severity was tracked weekly using the SPIN (Figure 5).

By the end of the 4-week intervention period, participants in RCT #1 with access to the Alena app saw a significantly greater reduction in SPIN (-9.83 ± 12.80) compared to the waitlist control group (-4.13 ± 11.59, t90 = -2.23, p_FDR_ = .037, Cohen’s d = 0.47, Figure 5A, B). Additionally, 51% percent of the intervention group showed a “clinically significant improvement” in social anxiety (≥ 10 point reduction), compared to only 22% of the control group (χ^2^ = 6.252, p = .012, Figure 5C). The percentage of participants reaching subclinical levels of social anxiety symptoms (SPIN ≤ 19) was not significantly different between the two groups (intervention: 19.15%, waitlist: 6.67%, χ^2^ = 2.153, p = .142).

At follow-up after six weeks, SPIN in the intervention group remained stable compared to the end of the intervention (0.05 ± 6.74), while the waitlist control group saw a reduction of 2.71 ± 6.10 points (t_90_ = 1.97, p_FDR_ = .052, Cohen’s d = 0.43, Figure 5A). This might be due to the fact that the waitlist control group had received access to the Alena app and 8% were using it, whereas the intervention group no longer had access to the Alena app. The difference in the number of participants showing a significant reduction of SPIN was no longer significantly different between the two groups at this timepoint (intervention: 46.6%, waitlist: 34.1%, χ^2^ = 1.522, p = .467), and neither was the difference in reliable recovery between groups (intervention: 17.78%, waitlist: 9.09%, χ^2^ = 0.791, p = .374).

These effects were broadly replicated in RCT #2. The intervention group showed a significantly larger reduction in SPIN scores (-12.89 ± 13.87) compared to the waitlist control group (-7.48 ± 12.24) by the end of the intervention period at eight weeks (t_227_ >= - 3.13, p_FDR_ = .008, Cohen’s d = 0.42, Figure 5D, E). A significantly larger proportion of the intervention group showed a clinically significant improvement in social anxiety compared to the control group (intervention: 60%, control 47.9%, χ^2^ = 12.908, p = .002, Figure 5F) and a larger group of participants in the intervention group reached subclinical levels of social anxiety symptoms by the end of the 8-week intervention (intervention: 21.9%, control: 10.48%, χ^2^ = 4.769, p = .006).

The effects in RCT #2 persisted at the week 12 follow-up, even though participants in neither group had access to the Alena app during this time. Indeed, participants in the intervention group continued to show a reduction in SPIN scores compared to the end of the intervention period (intervention: -2.39 ± 6.15, control: -0.29 ± 6.41, t_227_ = -2.48, p_FDR_ = 0.028, Cohen’s d = 0.33, Figure 5D). A larger proportion of participants assigned to the intervention group showed a clinically significant improvement in social anxiety (≥ 10 point reduction, intervention: 62.5%, control: 33.1%, χ^2^ = 17.008, p < .001) and they were 2.7 times more likely to have recovered (26.92%) than waitlist participants (11.57%, χ^2^ = 7.701, p = .006).

Lastly, a linear mixed-effects regression analysis on SPIN over the intervention period (including baseline), modulated by group (intervention vs waitlist) and controlling for age, sex (RCT #2 only), and the plateau effect of SPIN over time (week^2^) revealed a highly significant main effect of week (RCT #1: ß = -3.149 ± 0.398, t = -7.922, p <.001; RCT #2: ß = -3.388 ± 0.249, t = -13.589, p < .001), and group x week interaction in both RCTs (RCT #1: ß = 1.691 ± 0.566, t = 2.99, p = 0.003; RCT #2: ß = 1.588 ± 0.340, t = 4.669, p < .001), suggesting that SPIN scores declined in both groups, but this decline was significantly steeper in the intervention group (see Supplementary Material Table 1 for details).

Overall, these results suggest that having access to the Alena app significantly reduced social anxiety symptoms beyond the decrease observed in the waitlist control group. Furthermore, both RCTs show that this improvement persists over time, suggesting a lasting impact of the Alena app.

### Daily functioning

The impact of Alena on daily functioning was measured by the WSAS total scores. At baseline, all participants reported significant functional impairment with a median WSAS score of 19 across all participants.

RCT #1: By the end of the intervention, the intervention group showed a greater average reduction in WSAS scores (-4.53 ± 6.02) compared to the control group (-2.07 ± 5.71), although the difference was not statistically significant after adjusting for multiple comparisons (t90 = -2.01, p_FDR_ = .073, Cohen’s d = 0.42, Figure 7A, B). At the 2-week follow-up, the control group experienced a slight improvement (-1.24 ± 2.77), while the intervention group’s scores slightly worsened (1.05 ± 5.06), reaching statistical significance (t_82_ = 2.57, p_FDR_ = .048, Cohen’s d = 0.56).

**Figure 7.**
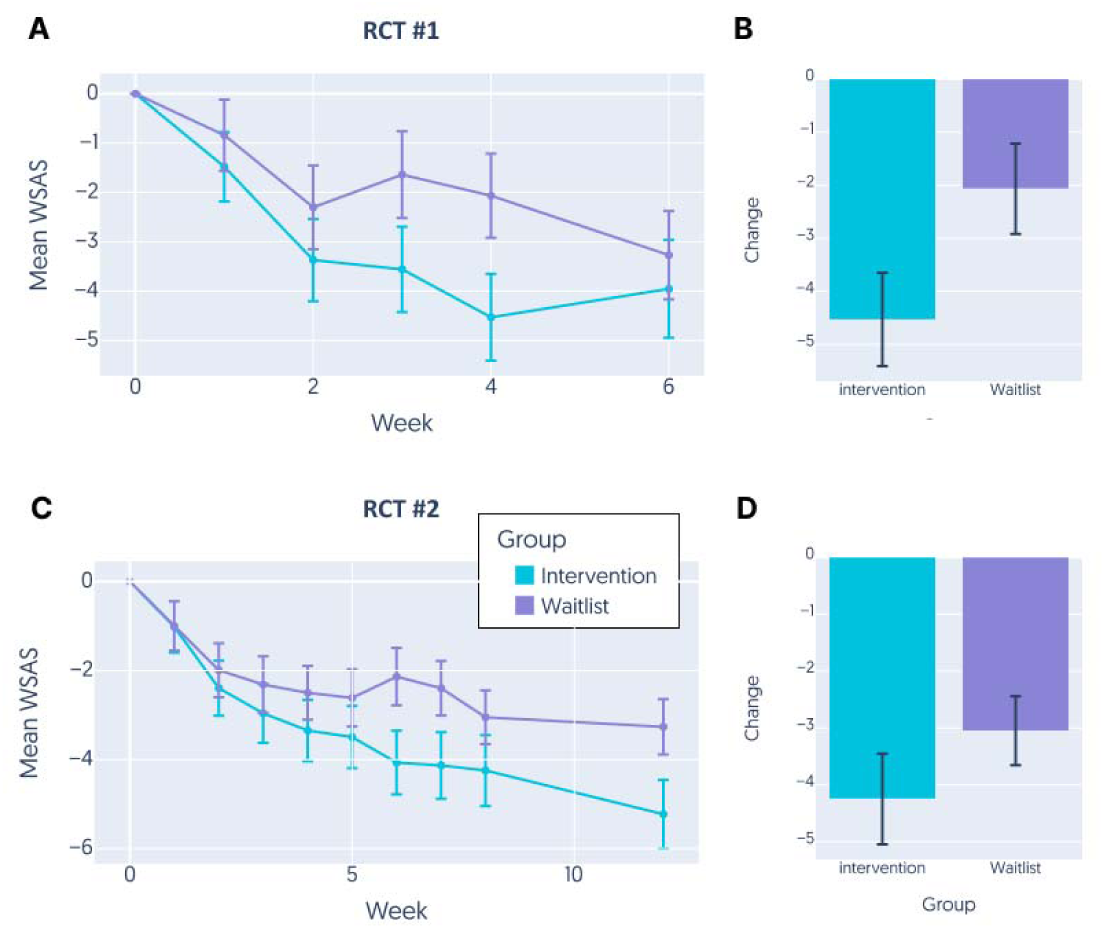
Improvement in daily functioning over time. **A** Mean change in WSAS score across participants in either the intervention (blue) or the waitlist control (purple) group each week relative to baseline assessment at week 0 in RCT #1. The final week is the follow-up assessment. All weeks in between constitute the intervention/waitlist period. **B** Total change in WSAS from week 0 to the final week of the intervention/waitlist period in RCT #1. **C** Mean change in WSAS score across participants in either the intervention (blue) or the waitlist control (purple) group each week relative to baseline assessment at week 0 in RCT #2. The final week is the follow-up assessment. All weeks in between constitute the intervention/waitlist period. **D** Total change in WSAS from week 0 to the final week of the intervention/waitlist period in RCT #2. Error bars represent standard error of the mean.

RCT #2: Throughout the 8-week intervention period, both groups demonstrated improvements in WSAS scores, with the intervention group seeing a slightly larger average reduction (-4.25 ± 8.19) than the control group (-3.05 ± 6.74), though the difference was not significant (t_227_ = -1.22, p_FDR_ = .225, Cohen’s d = 0.16). At the week 12 follow-up, changes in WSAS scores were minimal and not significantly different between the intervention (-1.04 ± 4.36) and waitlist control groups (0.08 ± 4.22, t_227_ = - 1.93, p_FDR_ = .073, Cohen’s d = 0.26)

In an exploratory analysis, a linear mixed-effects regression model analyzed the WSAS scores over time, adjusting for baseline values, group, age, and sex (in RCT #2 only), along with a quadratic time effect (week^2^). The analysis revealed a significant main effect of time, indicating that WSAS scores generally declined over the study period (RCT #1: ß = -1.572 ± 0.237, t = -6.629, p <.001; RCT #2: ß = -1.466 ± 0.152, t = -9.636, p < .001). The group by time interaction was also significant, suggesting that the rate of decline in WSAS scores was steeper in the intervention group compared to the control group (RCT #1: ß = 0.896 ± 0.337, t = 2.657, p = 0.008; RCT #2: ß = 0.756 ± 0.207, t = 3.646, p < .001, see Supplementary Material Table 2 for details).

The data indicate that the Alena app has a positive, though variable, impact on reducing functional impairments associated with social anxiety as measured by the WSAS. The intervention group generally showed a greater improvement in daily functioning across both RCTs, particularly notable given the significant interaction effects in the mixed-effects models.

## Discussion

### Principal results

The findings from our two randomized controlled trials provide evidence for the efficacy, safety, and acceptability of an online-only, modularized iCBT program for SAD in the form of a smartphone app (alena.com).

### Safety and acceptability

There were no indications that the online delivery of the interventions were unsafe. There was no increase in the severity or frequency of adverse events in the intervention group compared to the waitlist control group. The treatment included steps such as exposure which is crucial for therapeutic benefit but necessarily induces discomfort and could in principle be unsafe. The interventions made this very explicit, and provided instructions on how to engage in exposure safely. Hence, exposure can be delivered safely online without therapist involvement. This finding is also consistent with a recent meta-analysis that found that internet-based cognitive behavioral therapy (iCBT) is generally safer than control conditions [38].

### Acceptability

During the design of the app, detailed attention was paid to user design, including an appealing visual design, easy-to-use interface, bite-sized therapeutic content that was adapted to the participant population by providing relevant and normalizing examples. Overall, this led to high acceptability scores and positive user feedback. The high completion rate of the program (users completed 84-91% of material in the app) suggests that these features helped effectively engage users, encouraging consistent participation and adherence to the treatment protocol. As such, the results speak to the importance of careful user interface design for online interventions.

### Efficacy

The app demonstrated robust efficacy across both an unselected adult online sample, and in a sample of women aged 18-35. The latter group is particularly important given the high prevalence of social anxiety in younger women. In both groups, the positive treatment effects persisted after the treatment period ended, indicating at least a short-term maintenance of the benefits.

## Limitations

There are several important limitations to consider. The first is the short follow-up time. During this time, control group participants in RCT #1 also received the active treatment. Given that SAD is often viewed as a chronic disorder, it is important to determine whether the treatment has sustained effects over a longer period.

The second limitation concerns the sample, which was limited to active user of the online recruitment website prolific.com. Future studies should aim to include larger, more diverse samples. It is in particular not possible to generalize findings from a self-selected online panel of volunteers to a clinical sample. As such, the applicability of our results to real-world clinical populations will need to be assessed.

Third, we used a waitlist control as opposed to an active control condition as the comparison group in our study. Active controls, such as psychological placebos, often result in smaller effect sizes compared to passive controls [39,40]. This can be due to the fact that participants in the waitlist control group were aware of their status, which can lead to expectancy effects and other non-specific factors influencing outcomes. However, our primary goal was to establish the initial safety, acceptability and efficacy of the Alena app. A waitlist control allows for a clear comparison of the treatment effect without the confounding influence of another active intervention. Future studies should address this limitation by including active control groups to further validate the efficacy of the Alena app and mitigate potential biases. A direct comparison of digital interventions like Alena with their face-to-face therapy counterparts would provide valuable insights into the relative strengths and limitations of digital therapy, helping to refine these tools and better integrate them into mainstream mental health care. This direct comparison would also aid in identifying specific patient profiles that may benefit more from digital or traditional therapy modalities.

Fourth, although the therapy content is modularized, the current studies the delivered interventions in a standardized fashion, with modules released in a fixed order.

Lastly, we cannot rule out that the efficacy in RCT #2 is due to engagement with the Recharge and Community content that participants in the intervention group had access to throughout the duration of the trial. However, participants in RCT #1 did not have access to this content and effect sizes are comparable across the two trials. This suggests that the effects in RCT #2 can at least not be solely attributed to the additional content but likely result from the therapeutic interventions.

## Comparison with Prior Work

The efficacy of the Alena app either matches [41,42] or surpasses [7] that observed by previous digital interventions for social anxiety, although interventions including support from a human therapist can show enhanced effects [43]. In part, this probably reflects effective interface design driving engagement [44]. The efficacy seen here appears broadly comparable to in-person NHS Talking Therapy outcomes. In 2022, improvement rates of 67.1% and recovery rates of 36.4% for social phobia disorder treated with CBT were reported by NHS Talking Therapies [45]. This is similar to the observed improvement rates of 65% and recovery rates of 27% in RCT #2. The NHS also reports an average reduction in WSAS of 5.8 points, which is similar to the 5.2 point reduction we observed in RCT #2. Although it is not possible to compare clinical and the self-selected online samples, these comparisons encourage the examination of the app efficacy in clinical settings given the substantial cost-effectiveness and scalability nature of a smartphone app.

### Future directions

Here, iCBT retained efficacy despite the modularized format and the absence of a therapist intervention. This opens new doors for treating SAD by encouraging users to focus on specific cognitive and behavioral processes most relevant to their individual needs. The ability to tailor treatment plans based on individual profiles may ultimately help address the heterogeneity in symptom presentation and treatment response among individuals with SAD [29].

### Conclusions

In conclusion, the Alena app exemplifies the substantial potential of digital therapy for Social anxiety disorder, adapting a gold-standard model of CBT into a format that is safe, acceptable, effective and highly scalable. These findings pave the way for the development of accessible and tailored treatments for individuals with SAD and other mental health conditions. By advancing our understanding of how to implement modularized CBT in a digital format effectively, we move closer to achieving more personalized and effective mental health care for all.

## Data Availability

Data is not available.

## Acknowledgements

This study was sponsored and funded in full by Alena (Aya Technologies Ltd). We would like to thank Omar Reid for the technical support in setting up this study, and the many volunteers who provided generous feedback on the Alena App during its development.

## Abbreviations

AUDIT-C: Alcohol Use Disorders Identification Test for Consumption
BF01: Bayes Factor for the Null Hypothesis
CBT: Cognitive Behavioral Therapy
GP: General Practitioner
iCBT: Internet-delivered Cognitive Behavioral Therapy
ISO: International Organization for Standardization
NHS: National Health Service
RCT: Randomized Controlled Trial
SAD: Social Anxiety Disorder
SD: Standard Deviation
SPIN: Social Phobia Inventory
UCL: University College London
WSAS: Work and Social Adjustment Scale

## Contributions

The therapeutic content was designed by SL, with support by QJMH, SS, SSM and MMG. The app implementation was led by AL, with support by SL, MMG, SSM and SS. Conceptualization and study design were led by TM, MMG, QJMH, SL and AL. Data acquisition was undertaken by TM with support by SM and MMG. Data analysis was performed by JM, TM, MMG, PS and reviewed by QJMH. The manuscript was written by MMG and JM, with input from all authors.

