## Supplementary Material for "Safety and efficacy of a modular digital psychotherapy for social anxiety: A randomized controlled trial"

**Program outline**

RCT #1

| **Module** | **Format** | **Title** | **User-Facing Description** |
| --- | --- | --- | --- |
| Introduction | Interactive worksheet | What is social anxiety? | This chapter is an overview of social anxiety. You'll gain an  insight into your own symptoms and how to recognise them. |
|  | Interactive worksheet | How does Alena work? | Learn more about Alena's unique approach, which combines  evidence-based treatment with cutting-edge neuroscience  assessments. |
| Beliefs | Interactive worksheet | Map out your social anxiety | In this exercise you will learn how to use an evidence-based  tool called the Social Anxiety Map. |
|  | Interactive worksheet | Understand your thought patterns | In this exercise, you will learn about the three types of  negative thoughts which are common for social anxiety -  worries, beliefs and images. |
| Attention | Interactive worksheet | What is self-attention? | In this explainer, you will learn more about negative  self-attention and the role it plays in keeping social anxiety  going. |
|  | Interactive worksheet | Explore a new perspective | In this exercise you will work with a real situation from your  life and learn how to see it from more than one perspective. |
|  | Audio | Learn to shift your attention | In this guided meditation you will learn how to shift your  attention by focusing on different objects or sounds around  you. |
|  | Audio | Practice shifting attention | In this exercise, you will practise this new skill in a situation  you might encounter in the real world - giving a speech. |
| Avoidance | Interactive worksheet | What are safety behaviors? | Here you will learn more about avoidance and safety  behaviors, and the role they play in social anxiety. |
|  | Interactive worksheet | Prepare for real-life practice | This exercise will help you plan an experiment to drop a safety  behavior in a real-life social situation. |
|  | Interactive worksheet | Reflect on real-life practice | This reflection exercise will help you learn from your  experiment and identify the most helpful next steps for you, in  a non-judgemental and supportive way. |
| Rumination | Interactive worksheet | What is rumination? | In this explainer, you will learn more about rumination, how it  can spiral, and the role this plays in keeping social anxiety  going. |
|  | Interactive worksheet | Learn to stop negative thoughts | In this exercise you will learn an evidence-based technique to  help you break out of a negative thought spiral. |
|  | Interactive worksheet | Challenge your memory bias | In this exercise, you will learn a simple way to challenge any  tendency you may have to remember social events in a  negatively biased way. |

RCT #2

| **Module** | **Format** | **Title** | **User-Facing Description** |
| --- | --- | --- | --- |
| Introduction | Assessment | Check your social anxiety level | Get started with this assessment to measure your level of social anxiety. You will use it to track your progress over time. |
|  | Audio | What if it worked? | In this audio explainer, you will be guided to visualize what it would be like if working on your social anxiety with Alena worked. |
|  | Interactive worksheet | Are you ready? | In this audio explainer, you will be guided to visualize how it would be like if working on your social anxiety with Alena worked. |
|  | Interactive worksheet | Map out your social anxiety | In this exercise you will learn how to use an evidence-based tool called the Social Anxiety Map. |
|  | Audio | What is social anxiety? | This chapter is an overview of social anxiety. You'll gain an insight into your own symptoms and how to recognise them. |
|  | Audio | How does Alena work? | Learn more about Alena's unique approach, which combines evidence-based treatment with cutting-edge neuroscience assessments. |
|  | Interactive worksheet | Quiz - Test your knowledge | This quiz is designed to help you refresh your memory and consolidate the information you learned in this module. |
| Beliefs | Assessment | Assess your beliefs | Understand how your beliefs may be driving your social anxiety with this interactive assessment. |
|  | Audio | What are beliefs? | In this audio explainer, you will learn more about beliefs and the role they play in keeping social anxiety going. |
|  | Interactive worksheet | Balance a belief | Learn an evidence-based technique to challenge an unhelpful belief and create a more balanced one. |
|  | Interactive worksheet | Explore a new perspective | In this exercise you will work with a real situation from your life and learn how to see it from more than one perspective. |
|  | Audio | Applying it to your life | In this audio explainer you'll learn how to start setting up experiments to start changing your beliefs in real life. |
|  | Interactive worksheet | Put it into practice | In this exercise, you can set up your real-life experiment to challenge your beliefs, log your progress or journal about how it's going. |
| Attention | Assessment | Assess your attention | Play this fishing game where your goal is to work with a partner to catch as many fish as possible. |
|  | Audio | What is self-attention? | In this audio explainer, you will learn more about negative self-attention and the role it plays in keeping social anxiety going. |
|  | Audio | Visual attention training | In this guided meditation you will learn how to shift your attention by focusing on different objects around you. |
|  | Audio | Applying it to your life | This audio explainer will help you review what you have learned about self-attention and how to apply it to your life. |
|  | Interactive worksheet | Put it into practice | In this exercise, you can set up your real-life experiment to train your attention muscles, log your progress or journal about how it's going. Use this exercise to prepare guided experiments in real life situations. |
| Avoidance | Assessment | Assess your avoidance | In this assessment, you will be working with a partner and using your communication skills to gain tokens. |
|  | Audio | What is avoidance? | In this audio explainer, you will learn more about avoidance and safety behaviors, and the role they play in social anxiety. |
|  | Interactive worksheet | Identify a safety behaviour | In this exercise, you will reflect on one of your safety behaviors, and how it might be holding you back and keeping your anxiety going. |
|  | Interactive worksheet | Plan your experiment | This exercise will help you plan an experiment to drop a safety behavior in a real-life social situation. |
|  | Interactive worksheet | Reflect on your experiment | This reflection exercise will help you learn from your experiment and identify the most helpful next steps for you, in a non-judgemental and supportive way. |
|  | Audio | Applying it to your life | This audio explainer will help you celebrate your progress, review what you have learned about avoidance, and learn how to continue expanding your "stretch zone". |
|  | Interactive worksheet | Putting it into practice | In this exercise, you can set up experiments to drop your safety behaviors, log your progress or journal about how it's going. |
| Rumination | Assessment | Assess your rumination | You will play a code words game with others to uncover an impostor. Your goal is to guess the impostor and not get yourself caught! |
|  | Audio | What is rumination? | In this audio explainer, you will learn more about rumination, how it can spiral, and the role this plays in keeping social anxiety going. |
|  | Interactive worksheet | Learn to stop negative thoughts | In this exercise you will learn an evidence-based technique to help you break out of a negative thought spiral. |
|  | Interactive worksheet | Challenge your memory bias | In this exercise, you will learn a simple way to challenge any tendency you may have to remember social events in a negatively biased way. |
|  | Audio | Applying it to your life | This audio explainer will recap what you have learned about rumination and give you some helpful tips to for how to ruminate less. |
|  | Interactive worksheet | Put it into practice | Now you've worked on the cognitive side of rumination, put it into practice by testing it out in a real life situation, reflecting about it in a journal entry, or logging your progress. |

**Supplementary Table 1.** Linear mixed-effects regression analysis on the change in SPIN over the intervention period (including baseline but not including follow-up), modulated by group (intervention vs waitlist) and controlling for age, sex (RCT #2 only), and the plateau effect of SPIN over time (week^2^). Significant effects are highlighted in yellow.

Formula: SPIN ~ group × week + week^2^ + age + sex + (1|participant)

| **RCT** | **Effect** | **Estimate (β)** | **Standard error** | **t value** | **p value** | **d** |
| --- | --- | --- | --- | --- | --- | --- |
| RCT #1 | *Intercept* | *38.16* | *1.469* | *25.976* | *< 0.001 **** |  |
|  | Group | 2.002 | 2.059 | 0.972 | 0.333 | 0.17 |
|  | Week | -3.149 | 0.398 | -7.922 | < 0.001 *** | 0.27 |
|  | Week^2^ | 0.406 | 0.336 | 1.207 | 0.228 | 0.03 |
|  | Age | 1.408 | 1.03 | 1.367 | 0.175 | 0.12 |
|  | Group × Week | 1.691 | 0.566 | 2.99 | 0.003 | 0.15 |
| RCT #2 | *Intercept* | *36.044* | *1.226* | *29.399* | *< 0.001 **** |  |
|  | Group | 2.885 | 1.689 | 1.708 | 0.89 | 0.24 |
|  | Week | -3.388 | 0.249 | -13.589 | < 0.001*** | 0.28 |
|  | Week^2^ | 1.092 | 0.154 | 7.072 | < 0.001*** | 0.09 |
|  | Age | 0.019 | 0.677 | 0.028 | 0.978 | 0 |
|  | Sex | -1.788 | 1.988 | -0.899 | 0.369 | 0.15 |
|  | Group × Week | 1.588 | 0.340 | 4.669 | < 0.001*** | 0.13 |
|  | Group × Sex | -2.089 | 2.787 | -0.750 | 0.454 | 0.17 |
|  | Group × Week × Sex | 0.834 | 0.570 | 1.462 | 0.144 | 0.07 |

*d* is an estimate of effect size, where d=varrand . Variance Inflation Factor (VIF) < 2.5.

**Supplementary Table 2.** Linear mixed-effects regression analysis on the change in WSAS over the intervention period (including baseline but not including follow-up), modulated by group (intervention vs waitlist) and controlling for age, sex (RCT #2 only), and the plateau effect of SPIN over time (week^2^). Significant effects are highlighted in yellow.

Formula: WSAS ~ group × week + week^2^ + age + sex + (1|participant)

| **RCT** | **Effect** | **Estimate (β)** | **Standard error** | **t value** | **p value** | **d** |
| --- | --- | --- | --- | --- | --- | --- |
| RCT #1 | *Intercept* | *15.653* | *1.064* | *14.717* | *< 0.001 **** |  |
|  | Group | 2.093 | 1.504 | 1.391 | 0.167 | 0.26 |
|  | Week | -1.572 | 0.237 | -6.629 | < 0.001 *** | 0.19 |
|  | Week^2^ | 0.418 | 0.2 | 2.088 | 0.037 * | 0.05 |
|  | Age | -0.014 | 0.752 | -0.018 | 0.985 | 0 |
|  | Group × Week | 0.896 | 0.337 | 2.657 | 0.008 ** | 0.11 |
| RCT #2 | *Intercept* | *15.302* | *0.928* | *16.495* | *< 0.001**** |  |
|  | Group | 1.964 | 1.284 | 1.529 | 0.127 | 0.22 |
|  | Week | -1.466 | .152 | -9.636 | < 0.001*** | 0.17 |
|  | Week^2^ | 0.443 | 0.094 | 4.707 | < 0.001*** | 0.05 |
|  | Age | 0.497 | 0.514 | 0.966 | 0.335 | 0.06 |
|  | Sex | 1.116 | 1.505 | 0.742 | 0.459 | 0.13 |
|  | Group × Sex | -2.769 | 2.116 | -1.309 | 0.192 | 0.32 |
|  | Group × Week | 0.756 | 0.207 | 3.646 | < 0.001*** | 0.09 |
|  | Group × Week × Sex | -0.450 | 0.348 | -1.293 | 0.196 | 0.05 |

*d* is an estimate of effect size, where d=varrand . Variance Inflation Factor (VIF) < 2.5.

**Adverse health events reported by both groups**

**RCT #1**

**Treatment (week 1)**: Just had a chest infection following Covid. Follow this with GP who prescribed antibiotics.

**Treatment (week 2)**: Again chest infections being managed by antibiotics

**Treatment (week 2)**: I got covid for the first time and I was hospitalised because of it. I was exhausted and in pain all week.

**Treatment (week 3)**: Anxious throwing up

**Waitlist (week 1)**: I have been feeling worse than normal. I have been in a few social situations and I always feel like the odd one out.

**Waitlist (week 1)**: Not being interested in anything

**Waitlist (week 1)**: Doing physical tasks around the house that have caused slight damage to muscles and joints.

**Waitlist (week 2)**: I’m having severe stomach issues, symptoms of early menopause also, feeling more down and low in myself than usual

**Waitlist (week 2)**: Having to go up the hospital with my daughter. The place drains me mentally and takes me awhile to get back to my normal self, didn't help that we were there for hours

**Waitlist (week 2)**: Feeling numb

**Waitlist (week 3)**: Concussion

**Waitlist (week 3)**: Unable to sleep due to anxiety about new job

**Waitlist (week 3)**: Breathing problems

**Waitlist (week 3)**: Discussion about ADHD or Bipolar diagnosis

**Waitlist (week 4)**: Concussion recovery

**Waitlist (week 4)**: Anxiety and sleep disruption caused by stress about new job

**Waitlist (week 4)**: Daughter fell down the stairs had to go hospital back up there again this week due to her elbow not long coming out of plaster. Stressed and full of anxiety as she can't move her arm and I'm freaking out

**Waitlist (week 4)**: Really wanted to hurt myself. Urge

**RCT #2**

**Treatment (week 4)**: My physical tics are worse

**Treatment (week 5)**: Abnormal ecg

**Treatment (week 6)**: I was very worried

**Treatment (week 8)**: Feeling very stressed and vulnerable

**Treatment (week 8)**: Just been really lethargic and not wanting to do anything

**Treatment (week 8)**: I feel disappointed in myself as I have got stuck. It’s as likely to be very particular circumstances this week but I don’t feel like I can push forward with challenging myself. I haven’t had time to do the more reflective exercises this week which I would have liked to do and might have helped.

**Waitlist (week 4)**: Chest infection

**Waitlist (week 5)**: Anxiety and stress spiked, causing me to go on SSRIs

**Waitlist (week 5)**: Hallucinations, can't close my eyes without seeing unpleasant so really affecting my sleep

**Waitlist (week 5)**: Chest infection/ throat infection

**Waitlist (week 6)**: I can't finish things. My house is full of 'stuff' crafts I decided to take up but didn't keep up with, clothes that don't fit me, bags of I don't know what everywhere. I eat and drink far too much. I buy more clothes and brick a brac that I can't afford!

**Waitlist (week 6)**: Heightened mental health issues due to change in circumstances

**Waitlist (week 6)**: Worsening of depression and low mood

**Waitlist (week 7)**: I've turned to alcohol to try to shut out everything

**Waitlist (week 8)**: I have had to go for blood tests to do with my epilepsy and I'm concerned about the results that I don't know at the present

**Waitlist (week 8)**: I had a terribly acute sense of being overwhelmed and powerless.

**Waitlist (week 8)**: Breathing difficulty
